# Side Effects of COVID-19 Vaccines and Perceptions about COVID-19 and Its Vaccines in Bangladesh

**DOI:** 10.1101/2022.01.31.22270172

**Authors:** Md Mohsin, Sultan Mahmud, Ashraf Uddin Mian, Prottay Hasan, Abdul Muyeed, Md. Taif Ali, Fee Faysal Ahmed, Ariful Islam, Maisha Maliha Rahman, Mahfuza Islam, Md Hasinur Rahaman Khan, M. Shafiqur Rahman

## Abstract

**Introduction:** One of the primary reasons for hesitancy in taking COVID-19 vaccines is the fear of side effects. This study primarily aims to inspect the potential side effects of the COVID-19 vaccines circulated in Bangladesh.

**Methods:** The study was a cross-sectional anonymous online survey conducted across Bangladesh. Data were collected from December 2 to December 26, 2021. The study included consenting (informed) Bangladeshi individuals aged 12 and above who had received at least one dose of the COVID-19 vaccines. Analyses were carried out through exploratory analysis, Chi-square test, and logistic regression.

**Results:** A total of 1,180 (males-63.89%, age 50 years or over-65.4%, rural-52.86%) vaccinated people participated in the study. Less than half of the participants (39.48%) reported at least one side effect after receiving their COVID-19 vaccine. Injection-site pain, fever, headache, redness/swelling at the injection site, and lethargy were the most commonly reported adverse effects, all of which were mild and lasted 1-3 days. Side effects were most prevalent (about 80%) among individuals who received Pfizer-BioNTech and Moderna vaccines and were least common among those who received Sinopharm and Sinovac vaccines (21%-28%). When compared to the Sinopharm vaccines, the OxfordAstraZeneca, Pfizer-BioNTech, and Moderna vaccines were 4.51 (95% CI: 2.53-8.04) times, 5.37 (95% CI: 2.57-11.22) times, and 4.28 (95% CI: 2.28-8.05) times likelier to produce side effects. Furthermore, males, those over 50 years old, urban dwellers, smokers, and those with underlying health issues had a considerably increased risk of developing side effects. A lack of confidence in vaccines’ efficacy and a substantial level of hesitancy in allowing children (age five years or over) and older people (70 years or over) to receive COVID-19 vaccines were also observed.

**Conclusion:** Side effects of COVID-19 vaccines are minimal, demonstrating their safety. Further studies are required to establish the efficacy of the vaccines.

**What is already known?:** Significant COVID-19 vaccine hesitancy has been observed globally, mainly due to vaccine safety and efficacy concerns. Until now, most of the data on COVID-19 vaccine safety and efficacy have been published in manufacturer-funded trials that adhere to regulatory criteria and are monitored by third parties. A lack of independent studies on vaccine safety may have a detrimental effect on vaccine acceptance, which should be intensified to combat the deadly virus.

**What are the new findings?:** After receiving the COVID-19 vaccines, less than half of the subjects experienced at least one side effect. The side effects were mild and regular and lasted 1-3 days only. Side effects were most prevalent among individuals who received Pfizer-BioNTech and Moderna vaccines and were least common among those who received Sinopharm and Sinovac vaccines. A lack of confidence in vaccines’ efficacy and a substantial level of hesitancy in allowing children (age five years or over) and older people (70 years or over) to receive COVID-19 vaccines were also observed.

**What do the new findings imply?:** Side effects of COVID-19 vaccines are minimal and regular, demonstrating their safety. Efforts should be made to disseminate such findings among marginalized people worldwide who showed greater vaccine hesitancy.

## 1 Introduction

The COVID-19 pandemic has spread to every country on the planet, infecting nearly 270 million people and killing 5.4 million people as of December 11, 2021 [1]. COVID-19’s advent has had a disastrous influence on worldwide healthcare systems, with consequences in every facet of human life; leaving in its wake economic, familial, and mental health crises [2–4]. As a result, governments worldwide implemented border closures, travel bans, and quarantine protocols to stop the virus from spreading [4]. Unfortunately, the pandemic continues to wreak havoc across the globe.

Vaccines are thought to help the human body develop a long-lasting immune response to fight infectious diseases effectively. Indeed, vaccination prevents about 2–3 million deaths each year [5]. Vaccine development, however, is not the final word in eradicating such a widespread and deadly disease [4]. Vaccine hesitancy has been and continues to be a significant threat to mass vaccination [6]. It is a growing public health problem fueled by misconceptions about vaccine safety and effectiveness [7–9]. The most common cause of vaccine hesitancy (VH) among demographic groups in the United Kingdom (UK) was an aversion to vaccinations’ potential side effects, according to recent national research [10]. This conclusion was supported in the context of COVID-19 vaccinations, where fear of adverse effects was the most common reason for healthcare personnel and students in Poland declining to accept the vaccine [11,12]. As a result, a systematic evaluation of VH-fighting tactics found that increasing public awareness of vaccine effectiveness and transparency about side effects is critical for increasing vaccine uptake [13].

We are in a rapid infection spread caused by the virus (SARS-CoV-2) since it continuously mutates and spreads rapidly [14]. We have already seen 12 variants as of today, the Delta and the latest Omicron [15] being considered the most contagious [16]. In this evolving situation, widespread immunization is critical to preventing the catastrophic COVID-19 pandemic. Therefore, the Bangladesh government started a vaccination program from the beginning of 2021 and approved seven vaccines for mass immunization in Bangladesh. They are Covishield (Oxford/AstraZeneca), Pfizer/BioNTech (BNT162b2), Moderna (mRNA-1273), Johson & Johnson (Ad26.COV2.S), Sinopharm (BBIBP-CorV), Sinovac (CoronaVac), and Sputnik-V (Gamaleya) [17].

Bangladesh is a highly-populated country, and most of the people live in rural areas where misinformation and rumors are common. Hence, widespread ignorance, misinformation, and a lack of understanding concerning COVID-19 vaccines have persisted among the general public in Bangladesh since the start of the COVID-19 pandemic [18]. A significant VH has been found in Bangladesh per a cross-sectional study conducted in February 2021.

According to the study, among people willing to take a COVID-19 vaccine (61%), only 35% were willing to take a vaccine immediately if available [4]. The main reason for the unwillingness was doubts regarding the vaccines’ safety and efficacy [4]. As of December 11, 2021, only 25% of Bangladesh’s 160 million people have been fully vaccinated [19]. Vaccine hesitancy might play a vital role in low vaccine uptake in Bangladesh. Until now, most of the data on COVID-19 vaccine safety and efficacy have been published in manufacturer-funded trials that adhere to regulatory criteria and are monitored by third parties [20]. A lack of independent studies on vaccine safety may have a detrimental effect on vaccine acceptance, which must be intensified to combat the spread of the virus. A few studies have already examined a specific vaccine’s side effects. However, no studies have been found in the literature that examined most of the approved COVID-19 vaccines’ side effects. Here, side effects refer to any common or severe effects such as pain and redness/swelling at the injection site, fever, headache, etc., that occur after taking a COVID-19 vaccine.

The objectives of this study were to inspect the side effects of the circulated COVID-19 vaccines in Bangladesh, identify potential risk factors of the vaccine side effects, and explore the perceptions about COVID-19 and its vaccines among general people in general people Bangladesh.

## 2 Methods

### 2.1 Study Design

The study is based on a cross-sectional anonymous online survey conducted across Bangladesh and sought to shine a light upon the prevalence of the side effects of a range of COVID-19 vaccines on the Bangladeshi population. Participants in this survey had to be at least 12 years old and take at least one dose of a COVID-19 vaccine in Bangladesh. At the outset, a section described the study’s aim, the questionnaire’s concept, assurances regarding respondents’ confidentiality, and the study’s voluntary nature. Additionally, it was indicated that participants could omit a question if it appeared to be sensitive. The online survey began with the respondents’ informed consent and eligibility verification. After completing the survey, participants were also asked to share the survey link with their connections.

Additionally, participants/recipients of the survey link were asked to assist individuals in their families (younger and older) filling out the questionnaire who don’t have internet or social media access. The study questionnaire was prepared in English (see online supplemental questionnaire) and then translated into Bangla. Several experts and pilot surveys were used to validate the questionnaire. A link to an online survey (SurveyCTO) was shared on social media among thousands of presumably vaccinated people in Bangladesh (FB, Messenger, WhatsApp, and Electronic Email).

### 2.2 Timeframe and Inclusion Criteria

Data collection was done in Bangladesh from December 2 to December 26, 2021, using a web-based anonymous survey. Consenting Bangladeshi individuals of or over 12 years who received at least one dose of a COVID-19 vaccine were eligible for inclusion in the study.

### 2.3 Sample Size and Actual Response

A previous study [14] shows that 57% of general people had experienced the side effects of the COVID-19 vaccine. So then, the required minimum sample size is 501 calculated using the formula *SS=(Z*^*2*^*\*P(1-P)/*α ^*2*^*) *def*NR* where *Z*=1.96 at 95% confidence level, prevalence (*p*=0.5) of side effect of COVID-19 vaccines, the margin of error (α = 0.03); design effect (*def*= 1.6) for sampling variation; social media response rate from a previous study 70% [21].

### 2.4 Instruments

The study questionnaire was developed through an extensive literature review of similar studies with an eye on the context of Bangladesh. The survey comprised of questions regarding (i) Demographics (ii) COVID-19 Vaccination(s) Taken (iii) Underlying Health Conditions (iv) Side Effects of COVID-19 Vaccines (v) Knowledge of and Attitudes towards COVID-19 and its Vaccines. A panel of six experts with expertise in COVID-19 research and survey design were formed to review the questionnaire draft and assess its content validity. With ratings from the six experts, we computed a mean content validity index for items (I-CVI) of 0.946. According to Polit and Beck, with ratings from six or more experts, a mean I-CVI>=0.78 is considered good [22]. To estimate the instrument’s internal consistency, we used Chronbach’s Alpha statistic, and we found an Alpha score of 0.71, which is acceptable [23].

### 2.5 Consent and Ethical Considerations

The study leads with explicit declarations of anonymity by design, objectives, and voluntary nature. Participants could skip any questions if they found one uncomfortable to answer. The study was approved by the Ethical Review Committee, Faculty of Biological Science and Technology, Jashore University of Science and Technology, Jashore-7408, Bangladesh (Ref: ERC/FBST/JUST/2022-97).

### 2.6 Statistical Analysis

The exploratory analysis (bivariate analysis, frequencies analysis, means, graphs, etc.) was conducted to inspect the raw data. The Chi-square test was performed to determine the correlation between demographic factors and vaccines’ side effects. The multivariate logistic regression was used to identify the responsible factors for the intensity of the vaccines’ side effects among general people. The covariates that showed statistically significant association with vaccine side effects at 20% level of significance in the Chi-square test were included in the logistic regression model. We used Statistical software Stata (version 16) and R (version 4.1.2) to analyze and create graphs.

### 2.7 Patient and Public Involvement

This study did not include any patients. It was an online-based, voluntary, and anonymous study that collected data from general people aged 12 years or over who took at least one dose of a COVID-19 vaccine in Bangladesh. A comprehensive consent statement was included at the beginning of the survey describing the study’s objectives, nature, types of questions to be asked, skipping options, etc. The consent also assured that the data would be used in a combined form only for research purposes.

## 3 Results

### 3.1 Background Characteristics and Vaccine Prevalence

Table 1 describes the background characteristics of those who took part in the survey. The respondents tended to be male (63.89%) and over the age of 50 (65.40%). Most respondents indicated that they were married (65.40%). Respondents were evenly split between urban (47.14%) and rural (52.86%) regions. The majority of respondents indicated having received the Sinopharm vaccine (66.50%), followed by Oxford/AstraZeneca (10.69%), Moderna (7.66%), and Pfizer-BioNTech (7.32%). However, only 1.60% of respondents received the Sinovac vaccine, and the remaining 6.23% did not know the name of the vaccine they had received. The Sinopharm vaccine was also distinctly more prevalent in rural areas than urban Bangladesh. OxfordAstraZeneca, Pfizer-BioNTech and Moderna vaccinations were mainly reported by respondents of urban areas Figure 1.

**Table 1:** Socio-demographic charactereistics of the respondents

| Varibale | Labels | %(N) |
| --- | --- | --- |
| Consent | Yes | 100.00 (1,180) |
| Gender | Male | 63.89 (759) |
|  | Female | 36.11 (429) |
| Age | 12 to 29 years | 7.58 (90) |
|  | 30 to 39 years | 10.77 (128) |
|  | 40 to 49 years | 16.25 (193) |
|  | 50 to 59 years | 24.58 (292) |
|  | 60 or over | 40.82 (485) |
| Marital status | Single | 30.05 (357) |
|  | Married | 65.40 (777) |
|  | Other | 4.55 (54) |
| Education | No formal education (Illiterate) | 15.24 (181) |
|  | Primary completed | 7.49 (89) |
|  | Higher secondary (grade 6-10) | 9.85 (117) |
|  | SSC or equivalent (10th grade) | 11.11 (132) |
|  | HSC or equivalent (12th grade) | 12.71 (151) |
|  | Undergraduate (Hon's/MBBS/Degree/Technical) | 27.10 (322) |
|  | Graduate (Masters/PHD/Mphill) | 16.50 (196) |
| Income | 10,000-19,999 | 40.91 (486) |
|  | 20,000-29,999 | 14.73 (175) |
|  | 30,000-39,999 | 8.59 (102) |
|  | 40,000-49,999 | 6.06 (72) |
|  | 50,000-74,999 | 4.29 (51) |
|  | 75,000 or over | 4.46 (53) |
|  | Don't know | 20.96 (249) |
| Occupation | Small business (less than 5 employees) | 31.14 (370) |
|  | Large business (5 or more employees) | 1.77 (21) |
|  | Day laborer/Rickshaw/Van/Auto driver | 8.33 (99) |
|  | Motor vehicle driver | 1.01 (12) |
|  | Student | 22.05 (262) |
|  | Housewife | 22.90 (272) |
|  | Unemployed | 5.89 (70) |
|  | Retied/Disabled/Sick | 5.05 (60) |
|  | Other | 1.85 (22) |
| Religion | Islam | 92.00 (1,093) |
|  | Hinduism | 6.23 (74) |
|  | Christianity | 1.77 (21) |
| Region | Urban | 47.14 (560) |
|  | Rural | 52.86 (628) |
| Name of the vaccine | OxfordAstraZeneca | 10.69 (127) |
|  | Pfizer-BioNTech | 7.32 (87) |
|  | Moderna | 7.66 (87) |
|  | Sinopharm | 66.50 (790) |
|  | Sinovac | 1.60 (19) |
|  | Don't know the name | 6.23 (74) |
| Smoking status | No | 69.02 (820) |
|  | Yes | 30.98 (368) |
| Drink (Alcohol)/take illicit substances (Gaja/Yaba, etc.) | No | 94.53 (1,123) |
|  | Yes | 5.47 (65) |

Other: Freelancer, Researcher, Agriculture

**Figure 1:**
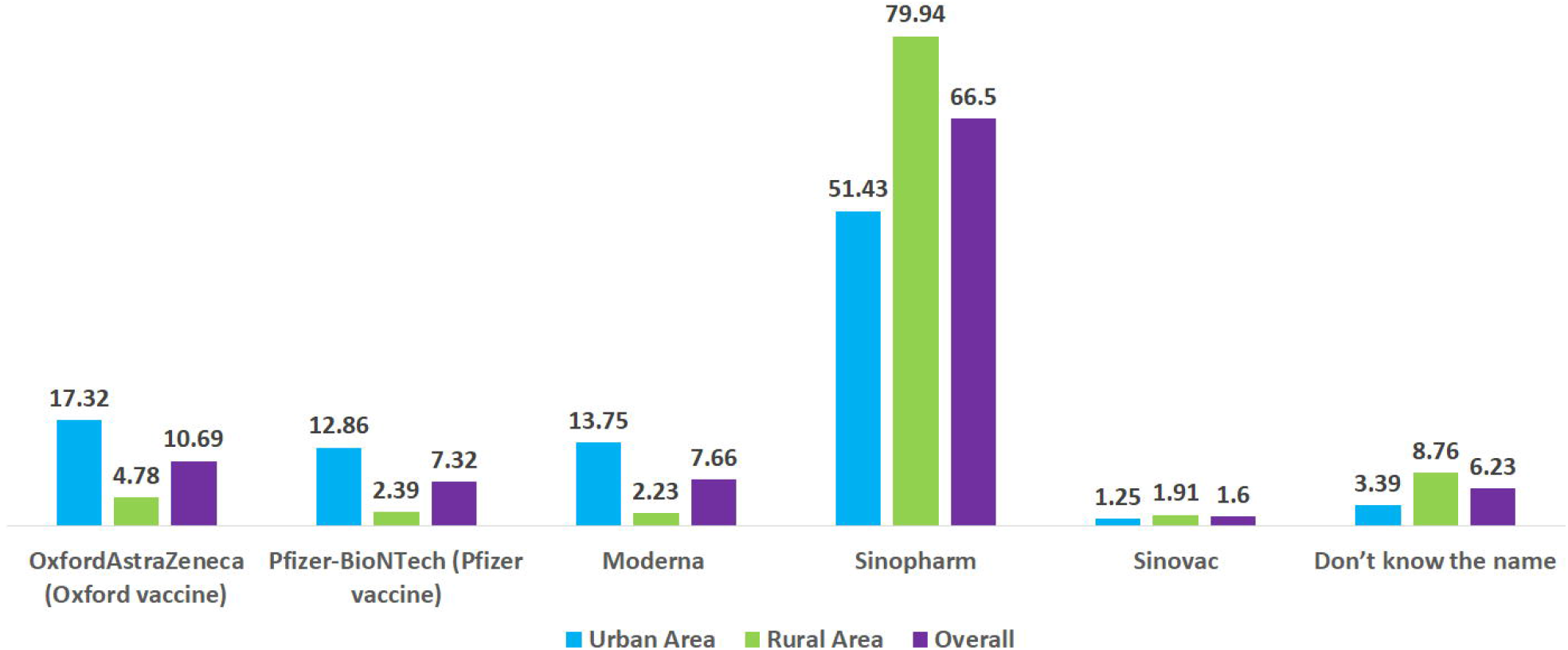
Distribution of vaccines over residence type

Respondents came from varied educational backgrounds—as measured by the highest degree obtained. While undergraduate-passed led with 27.10%, followed by graduate degree passed (16.50%), there were many without formal education (15.24%), HSC (level 12^th^)-passed (12.71%) or SSC (level 10^th^)-passed (11.11%). Respondents earned mainly in the 10,000-19,999 range (40.91%), but several (20.96%) indicated they don’t know, perhaps indicating reservations about disclosing income information. Respondents were most likely to be workers in a small business (31.14%) (large business counterparts stood at a much lower 1.77%), students (22.05%) or housewives (22.90%). The unemployed and retired/disabled/sick made up around 10.94% of respondents. Respondents’ religious profiles roughly tracked the Bangladeshi demographic statistic at 92.00% Muslim, 6.23% Hindu and 1.77% Christian.

Most respondents neither smoked (69.02%) nor, by an overwhelming majority, reported drinking or substance abuse (94.53%). Respondents reported underlying health conditions such as diabetes (8.98%), hypertension/High blood pressure (5.89%), severe allergies (5.22%), low blood pressure (5.81%) and chronic respiratory diseases (Pneumonia, Asthma, breathing issues) (4.71%) as described in Figure 2. A significantly smaller portion of the respondents has liver/kidney disease (1.18%), anemia (1.94%), heart disease/heart attack (2.69%) as well as obesity (2.86%).

**Figure 2:**
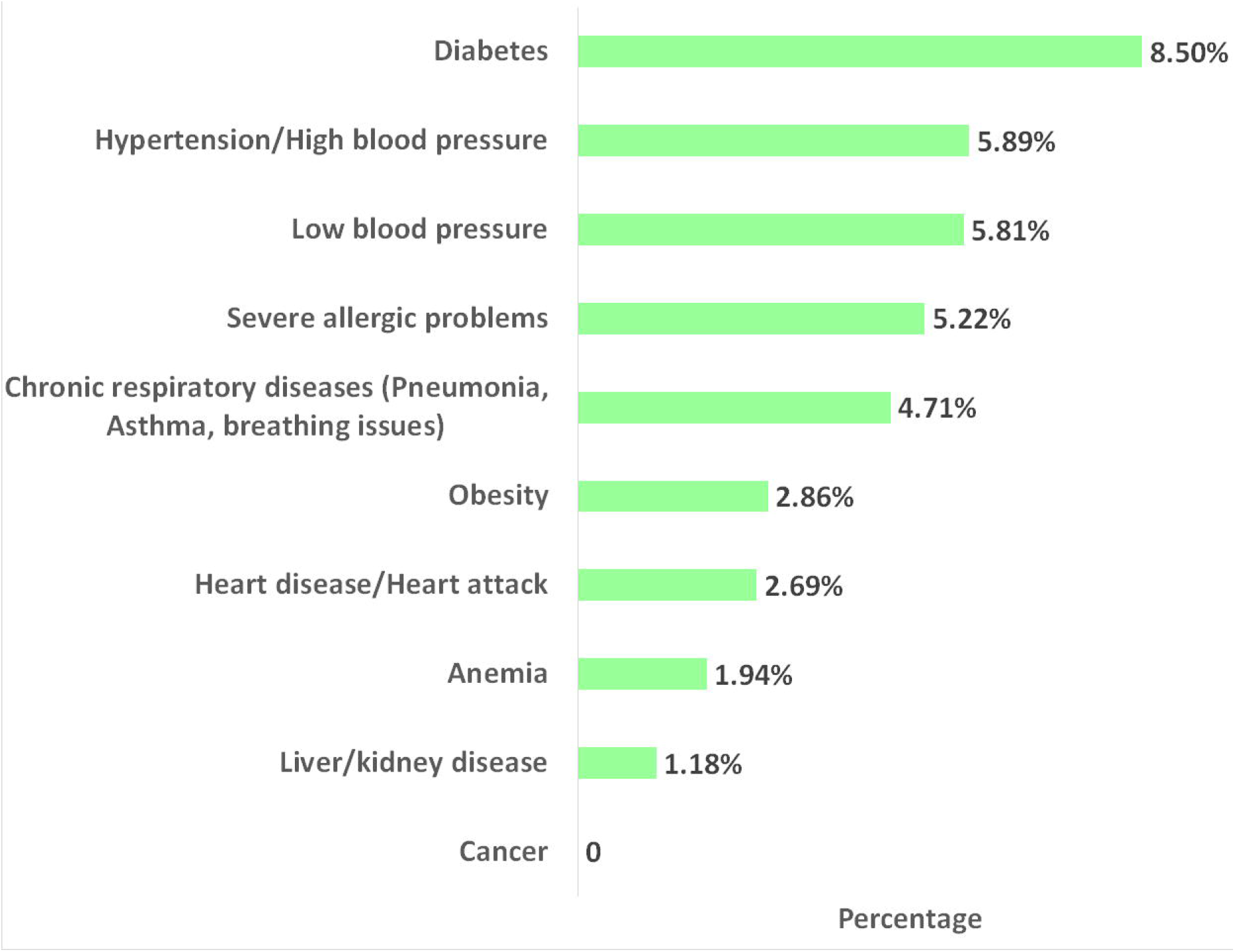
Distribution of underlying health conditions

### 3.2 Distribution of COVID-19 Vaccines’ Side Effects and their Severity

Overall, 39.48% of the participants experienced at least one side effect after receiving a COVID-19 vaccine in Bangladesh (Figure 3). The highest percentage (80.46%) of side effects were observed among people who received the Pfizer-BioNTech vaccine, and the second-highest prevalence of side effects (76.63%) was found among people who received Moderna, followed by 67.72% among people who took OxfordAstraZeneca vaccines (see Figure 4). The lowest percentage of side effects was found among people who received Sinopharm (28%.23) and Sinovac (21.05%) vaccines. Table 2 shows that respondents who faced side effects for taking the OxfordAstraZeneca vaccine, 86% of them had to take medicines. Most of them suffered from injection site pain (96.51%), fever (94.19%), headache (81.40%), and redness/swelling at the injection site (79%). Very few of them slept less (14.29%) and were anxious (3.49%). A large proportion of the respondents who took the Pfizer vaccine suffered from injection site pain (90%), fever (80%), and headache (74.29%). Likewise, among those who received the Moderna vaccine, 97%, 91%, 68.29% of participants suffered from injection site pain, fever and headache, respectively. More than 70% of the respondents who faced side effects for Pfizer and Moderna vaccines, took medicines. In contrast, only 9.87% of people took medicine who received Sinopharm and faced side effects. Moreover, around 50% to 70% of respondents who took the Sinovac vaccine mentioned having injection site pain, fever, or headache. Figure 5 shows the distribution of symptoms lasting duration (in terms of the number of days) across different COVID-19 vaccines. Psychological issues like less sleep and anxiety were more prevalent among those who took the OxfordAstraZeneca vaccine. However, symptoms durations were considerably short for those who received Sinopherm and Sinovac vaccines.

**Figure 3:**
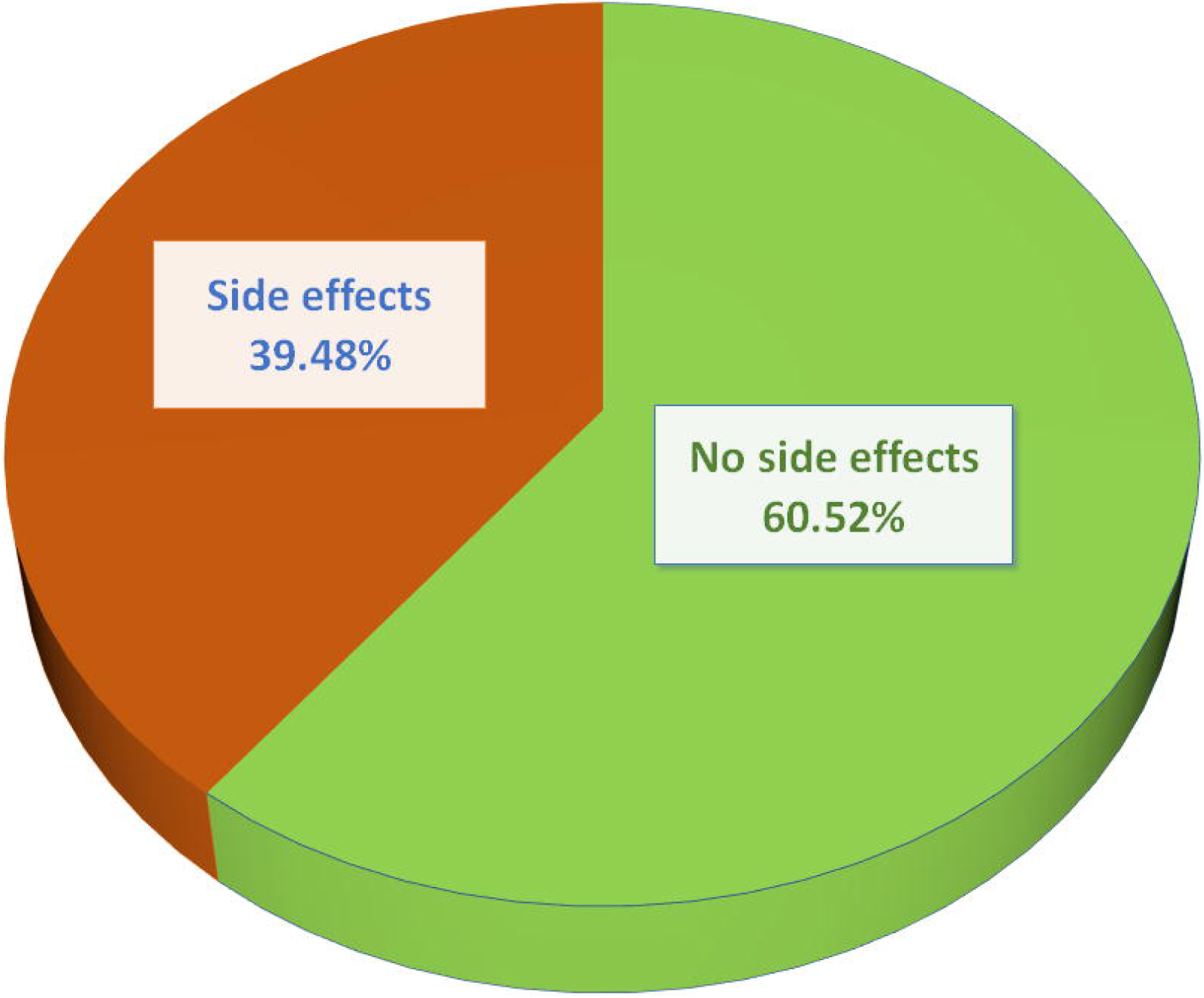
Overall side effects of COVID-19 vaccines among the general population in Bangladesh irrespective of vaccine type

**Figure 4:**
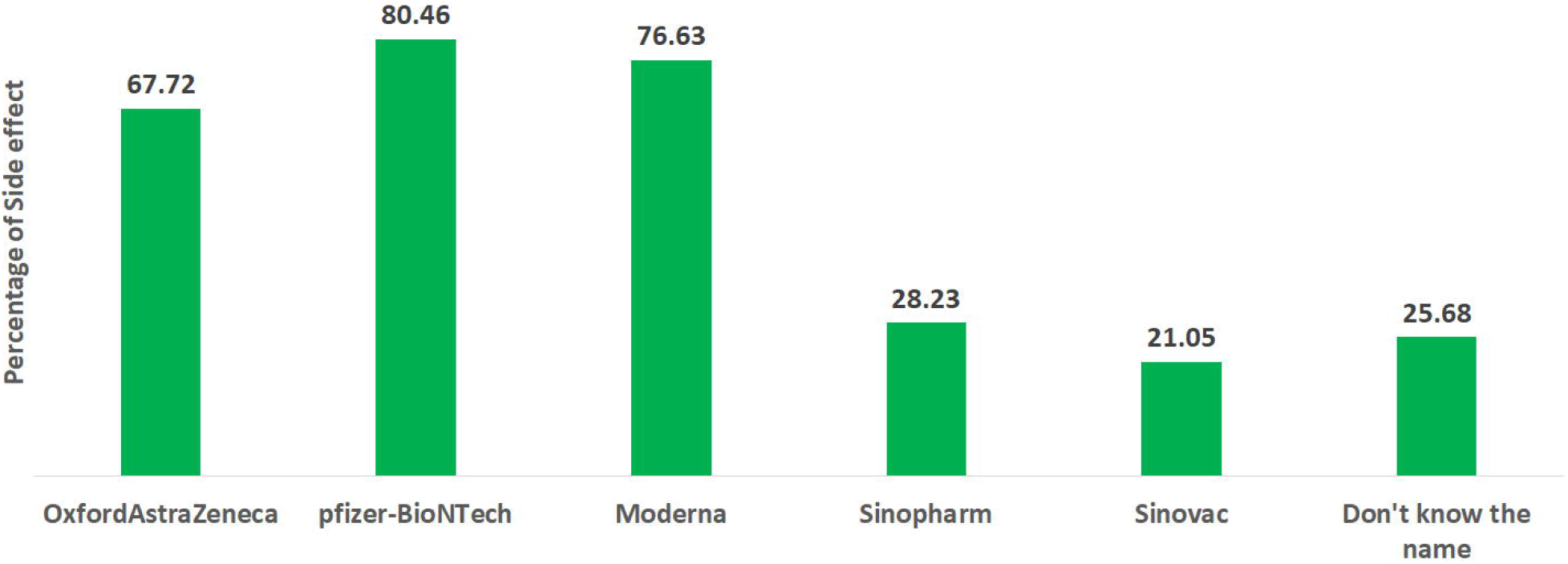
Percentage distribution of side effects of different COVID-19 vaccines among Bangladeshi people

**Figure 5:**
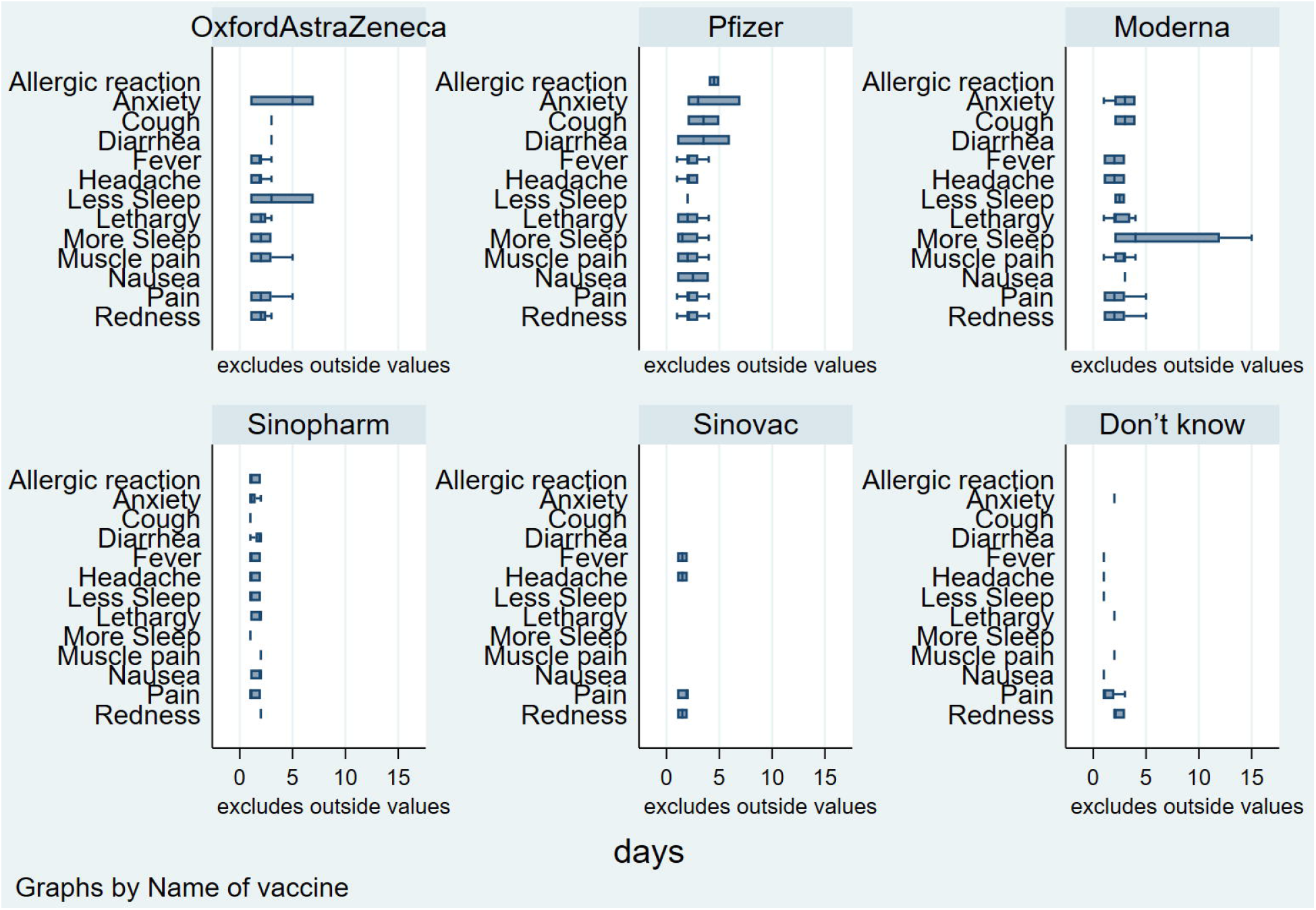
Distribution of symptoms duration (number of days) across different COVID-19 Vaccines

**Table 2:** Percentage distribution of side effects across different COVID-19 vaccines

| Name of the Vaccine | Symptoms | Frequency | Percentage |
| --- | --- | --- | --- |
| OxfordAstraZeneca | Total | 86 | 67.72 |
|  | Had to take medicine | 74 | 86.00 |
|  | Injection site pain | 83 | 96.51 |
|  | Redness/swelling at injection site | 68 | 79.07 |
|  | Fever | 81 | 94.19 |
|  | Headache | 70 | 81.40 |
|  | Lethargy | 12 | 13.95 |
|  | Nausea | 86 | 100.00 |
|  | Diarrhea | 1 | 1.16 |
|  | Cough | 1 | 1.16 |
|  | Muscle pain | 9 | 10.47 |
|  | Anxiety | 3 | 3.49 |
|  | Less Sleep | 3 | 14.29 |
|  | More Sleep | 2 | 2.33 |
| Pfizer-BioNTech | Total | 70 | 80.46 |
|  | Had to take medicine | 55 | 78.00 |
|  | Injection site pain | 63 | 90.00 |
|  | Redness/swelling at injection site | 50 | 71.43 |
|  | Fever | 56 | 80.00 |
|  | Headache | 52 | 74.29 |
|  | Lethargy | 13 | 18.57 |
|  | Nausea | 2 | 2.86 |
|  | Diarrhea | 2 | 2.86 |
|  | Cough | 2 | 2.86 |
|  | Allergic reaction | 2 | 2.86 |
|  | Muscle pain | 15 | 21.43 |
|  | Anxiety | 3 | 4.29 |
|  | Less Sleep | 7 | 25.93 |
|  | More Sleep | 4 | 5.71 |
| Moderna | Total | 67 | 73.63 |
|  | Had to take medicine | 52 | 77.61 |
|  | Injection site pain | 65 | 97.01 |
|  | Redness/swelling at injection site | 50 | 74.63 |
|  | Fever | 61 | 91.04 |
|  | Headache | 46 | 68.66 |
|  | Lethargy | 12 | 40.00 |
|  | Nausea | 1 | 1.49 |
|  | Cough | 3 | 4.48 |
|  | Muscle pain | 17 | 25.37 |
|  | Anxiety | 5 | 7.46 |
|  | Less Sleep | 2 | 6.67 |
|  | More Sleep | 6 | 8.96 |
| Sinopharm | Total | 223 | 28.23 |
|  | Had to take medicine | 22 | 9.87 |
|  | Injection site pain | 213 | 95.52 |
|  | Redness/swelling at injection site | 128 | 57.40 |
|  | Fever | 174 | 78.03 |
|  | Headache | 128 | 57.40 |
|  | Lethargy | 44 | 37.93 |
|  | Nausea | 6 | 2.69 |
|  | Diarrhea | 4 | 1.79 |
|  | Cough | 5 | 2.24 |
|  | Allergic reaction | 8 | 3.59 |
|  | Muscle pain | 29 | 13.00 |
|  | Anxiety | 16 | 7.17 |
|  | Less Sleep | 15 | 12.93 |
|  | More Sleep | 15 | 6.73 |
| Sinovac | Total | 4 | 21.05 |
|  | Had to take medicine | 3 | 75.00 |
|  | Injection site pain | 3 | 75.00 |
|  | Redness/swelling at injection site | 2 | 50.00 |
|  | Fever | 2 | 50.00 |
|  | Headache | 2 | 50.00 |
|  | Nausea | 4 | 100.00 |
|  | Less Sleep | 2 | 100.00 |
| Don't know name | Total | 19 | 25.68 |
|  | Had to take medicine | 2 | 10.53 |
|  | Injection site pain | 17 | 89.47 |
|  | Redness/swelling at injection site | 3 | 15.79 |
|  | Fever | 6 | 31.58 |
|  | Headache | 1 | 5.26 |
|  | Lethargy | 2 | 10.53 |
|  | Nausea | 1 | 5.26 |
|  | Muscle pain | 4 | 21.05 |
|  | Anxiety | 1 | 5.26 |
|  | Less Sleep | 1 | 5.26 |

### 3.3 Factors Associated with COVID-19 Vaccine Side Effects

The multivariate logistic regression seeks to identify influential factors for experiencing COVID-19 vaacine’s side effects. It is based on those factors which have a significant association with experiencing side effects at a 20% level of significance (see Table 2). The estimated parameters from logistic regression generally have been interpreted in terms of the odds ratio. The odds are defined as the probability of experiencing the event divided by the probability of not experiencing the event [24,25]. The odds ratios presented in **Table 3** with a 95% confidence interval indicate the odds of experiencing side effects in one particular group compared to odds of experiencing side effects in the reference group. The parameters are considered statistically significant at a 5% level of significance.

**Table 3:** Association between potential factors and COVID-19 vaccine side effects

| | | Side effect of COVID-19 vaccine | | $\chi^2$ | P-Value |
| --- | --- | --- | --- | --- | --- |
|  |  | Yes | No |  |  |
| Gender | Male | 50.20 (381) | 49.80 (378) | 93.76 | <0.001 |
|  | Female | 78.79 (338) | 21.21 (91) |  |  |
| Age | 12 to 29 years | 23.33 (21) | 76.67 (69) | 84.29 | <0.001 |
|  | 30 to 39 years | 19.53 (25) | 80.47 (103) |  |  |
|  | 40 to 49 years | 27.98 (54) | 72.02 (139) |  |  |
|  | 50 to 59 years | 36.99 (108) | 63.01 (184) |  |  |
|  | 60 or over | 53.81 (261) | 46.19 (224) |  |  |
| Marital status | Single | 56.02 (200) | 43.98 (157) | 60.13 | <0.001 |
|  | Married | 32.95 (256) | 67.05 (521) |  |  |
|  | Other(specify) | 24.07 (13) | 75.93 (41) |  |  |
| Education | No formal education (Illiterate) | 16.57 (30) | 83.43 (151) | 134.16 | <0.001 |
|  | Primary completed | 23.60 (21) | 76.40 (68) |  |  |
|  | Higher secondary (grade 6-10) | 23.08 (27) | 76.92 (90) |  |  |
|  | SSC or equivalent completed (10<sup>th</sup> grade) | 30.30 (40) | 69.70 (92) |  |  |
|  | HSC or equivalent passed (12<sup>th</sup> grade) | 37.09 (56) | 62.91 (95) |  |  |
|  | Undergraduate (Hon's/MBBS/Degree/Technical) | 55.59 (179) | 44.41 (143) |  |  |
|  | Graduate (Masters/PHD/Mphill) | 59.18 (116) | 40.82 (80) |  |  |
| Income | 10,000-19,999 | 34.77 (169) | 65.23 (317) | 112.09 | <0.001 |
|  | 20,000-29,999 | 45.14 (79) | 54.86 (96) |  |  |
|  | 30,000-39,999 | 66.67 (68) | 33.33 (34) |  |  |
|  | 40,000-49,999 | 44.44 (32) | 55.56 (40) |  |  |
|  | 50,000-74,999 | 54.90 (28) | 45.10 (23) |  |  |
|  | 75,000 or over | 77.36 (41) | 22.64 (12) |  |  |
|  | Don't know | 20.88 (52) | 79.12 (197) |  |  |
| Occupation | Small business (less than 5 employees) | 52.43 (194) | 47.57 (176) | 140.9 | <0.001 |
|  | Large business (5 or more employees) | 47.62 (10) | 52.38 (11) |  |  |
|  | Day laborer/Rickshaw/Van/Auto driver | 16.16 (16) | 83.84 (83) |  |  |
|  | Motor vehicle driver | 25.00 (3) | 75.00 (9) |  |  |
|  | Student | 54.96 (144) | 45.04 (118) |  |  |
|  | Housewife | 15.81 (43) | 84.19 (229) |  |  |
|  | Unemployed | 38.57 (27) | 61.43 (43) |  |  |
|  | Retied/Disabled/Sick | 41.67 (25) | 58.33 (35) |  |  |
|  | Other | 31.82 (7) | 68.18 (15) |  |  |
| Religion | Islam | 38.24 (418) | 61.76 (675) | 17.11 | <0.001 |
|  | Hinduism | 45.95 (34) | 54.05 (40) |  |  |
|  | Christianity | 80.95 (17) | 19.05 (4) |  |  |
| Region | Urban | 66.61 (373) | 33.39 (187) | 326.32 | <0.001 |
|  | Rural | 15.29 (96) | 84.71 (532) |  |  |
| Name of the vaccine | OxfordAstraZeneca | 67.72 (86) | 32.28 (41) | 198.4 | <0.001 |
|  | Pfizer-BioNTech | 80.46 (70) | 19.54 (17) |  |  |
|  | Moderna | 73.63 (67) | 26.37 (24) |  |  |
|  | Sinopharm | 28.23 (223) | 71.77 (567) |  |  |
|  | Sinovac | 21.05 (4) | 78.95 (15) |  |  |
|  | Don't know the name | 25.68 (19) | 74.32 (55) |  |  |
| Smoking status | No | 23.41 (192) | 76.59 (628) | 285.89 | <0.001 |
|  | Yes | 75.27 (277) | 24.73 (91) |  |  |
| Drink (Alcohol)/take illicit substances (Gaja/Yaba, etc.) | No | 37.40 (420) | 62.60 (703) | 37.1 | <0.001 |
|  | Yes | 75.38 (49) | 24.62 (16) |  |  |
| Diabetes | No | 37.90 (412) | 62.10 (675) | 13.29 | <0.001 |
|  | Yes | 56.44 (57) | 43.56 (44) |  |  |
| Heart disease/Heart attack | No | 39.19 (453) | 60.81 (703) | 1.52 | 0.22 |
|  | Yes | 50.00 (16) | 50.00 (16) |  |  |
| Hypertension/High blood pressure | No | 40.16 (449) | 59.84 (669) | 3.7 | 0.05 |
|  | Yes | 28.57 (20) | 71.43 (50) |  |  |
| Low blood pressure | No | 38.07 (426) | 61.93 (693) | 16 | <0.001 |
|  | Yes | 62.32 (43) | 37.68 (26) |  |  |
| Cancer | No | 39.48 (469) | 60.52 (719) | - | - |
|  | Yes | 0 (0) | 0 (0) |  |  |
| Obesity | No | 38.73 (447) | 61.27 (707) | 9.32 | 0.002 |
|  | Yes | 64.71 (22) | 35.29 (12) |  |  |
| Severe allergic problem | No | 37.48 (422) | 62.52 (704) | 36.13 | <0.001 |
|  | Yes | 75.81 (47) | 24.19 (15) |  |  |
| Chronic respiratory diseases (Pneumonia, Asthma, breathing issues) | No | 38.25 (433) | 61.75 (699) | 15.14 | <0.001 |
|  | Yes | 64.29 (36) | 35.71 (20) |  |  |
| Liver/Kidney disease | No | 39.35 (462) | 60.65 (712) | 0.66 | 0.42 |
|  | Yes | 50.00 (7) | 50.00 (7) |  |  |
| Anemia | No | 38.63 (450) | 61.37 (715) | 18.26 | <0.001 |
|  | Yes | 82.61 (19) | 17.39 (4) |  |  |

Table 4 displays the results of the logistic regression model. Vaccine side effects were significantly associated with types of COVID-19 vaccine. For example, the odds of having COVID-19 vaccine side effects among people who took the OxfordAstraZeneca vaccine were 4.51 (2.53-8.04) times higher than people who took the Sinopharm vaccine. Pfizer-BioNTech receivers showed 5.37 (2.57-11.22) times higher odds of side effects than Sinopharm receivers. Likewise, respondents vaccinated with Moderna experienced 4.28 (95% CI: 2.28-8.05) times higher side effects than those who took the Sinopharm vaccine.

**Table 4:**
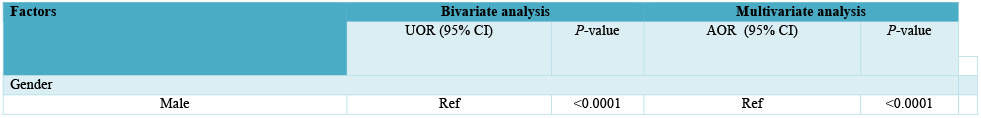
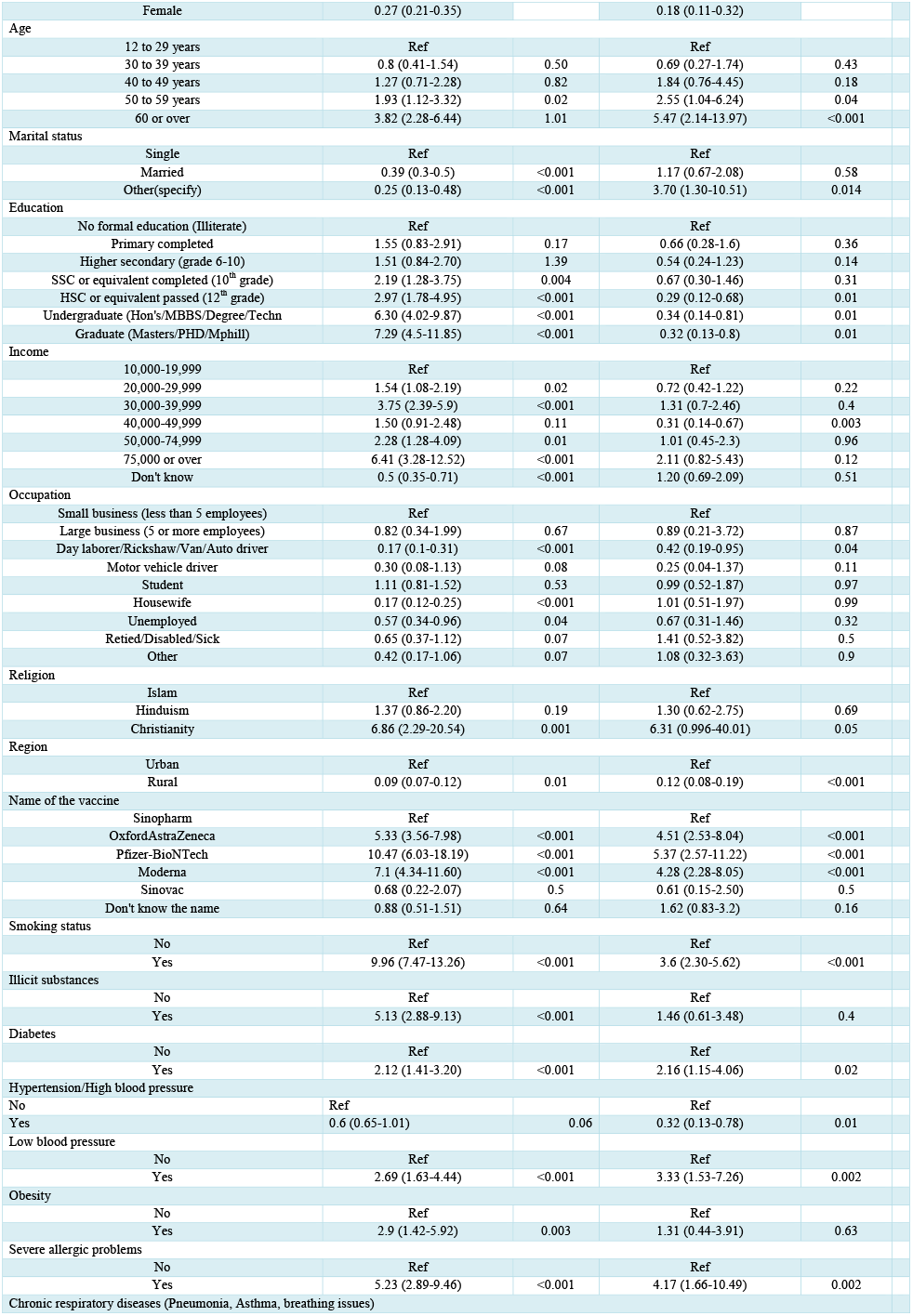
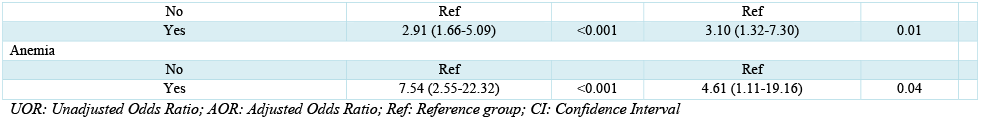
Factors associated with COVID-19 vaccine side effects

| Factors | Bivariate analysis |  | Multivariate analysis |  |
| --- | --- | --- | --- | --- |
|  | UOR (95% CI) | P-value | AOR (95% CI) | P-value |
| Gender |  |  |  |  |
| Male | Ref | <0.0001 | Ref | <0.0001 |
| Female | 0.27 (0.21-0.35) |  | 0.18 (0.11-0.32) |  |
| Age |  |  |  |  |
| 12 to 29 years | Ref |  | Ref |  |
| 30 to 39 years | 0.8 (0.41-1.54) | 0.50 | 0.69 (0.27-1.74) | 0.43 |
| 40 to 49 years | 1.27 (0.71-2.28) | 0.82 | 1.84 (0.76-4.45) | 0.18 |
| 50 to 59 years | 1.93 (1.12-3.32) | 0.02 | 2.55 (1.04-6.24) | 0.04 |
| 60 or over | 3.82 (2.28-6.44) | 1.01 | 5.47 (2.14-13.97) | <0.001 |
| Marital status |  |  |  |  |
| Single | Ref |  | Ref |  |
| Married | 0.39 (0.3-0.5) | <0.001 | 1.17 (0.67-2.08) | 0.58 |
| Other(specify) | 0.25 (0.13-0.48) | <0.001 | 3.70 (1.30-10.51) | 0.014 |
| Education |  |  |  |  |
| No formal education (Illiterate) | Ref |  | Ref |  |
| Primary completed | 1.55 (0.83-2.91) | 0.17 | 0.66 (0.28-1.6) | 0.36 |
| Higher secondary (grade 6-10) | 1.51 (0.84-2.70) | 1.39 | 0.54 (0.24-1.23) | 0.14 |
| SSC or equivalent completed (10<sup>th</sup> grade) | 2.19 (1.28-3.75) | 0.004 | 0.67 (0.30-1.46) | 0.31 |
| HSC or equivalent passed (12<sup>th</sup> grade) | 2.97 (1.78-4.95) | <0.001 | 0.29 (0.12-0.68) | 0.01 |
| Undergraduate (Hon's/MBBS/Degree/Techn | 6.30 (4.02-9.87) | <0.001 | 0.34 (0.14-0.81) | 0.01 |
| Graduate (Masters/PHD/Mphil) | 7.29 (4.5-11.85) | <0.001 | 0.32 (0.13-0.8) | 0.01 |
| Income |  |  |  |  |
| 10,000-19,999 | Ref |  | Ref |  |
| 20,000-29,999 | 1.54 (1.08-2.19) | 0.02 | 0.72 (0.42-1.22) | 0.22 |
| 30,000-39,999 | 3.75 (2.39-5.9) | <0.001 | 1.31 (0.7-2.46) | 0.4 |
| 40,000-49,999 | 1.50 (0.91-2.48) | 0.11 | 0.31 (0.14-0.67) | 0.003 |
| 50,000-74,999 | 2.28 (1.28-4.09) | 0.01 | 1.01 (0.45-2.3) | 0.96 |
| 75,000 or over | 6.41 (3.28-12.52) | <0.001 | 2.11 (0.82-5.43) | 0.12 |
| Don't know | 0.5 (0.35-0.71) | <0.001 | 1.20 (0.69-2.09) | 0.51 |
| Occupation |  |  |  |  |
| Small business (less than 5 employees) | Ref |  | Ref |  |
| Large business (5 or more employees) | 0.82 (0.34-1.99) | 0.67 | 0.89 (0.21-3.72) | 0.87 |
| Day laborer/Rickshaw/Van/Auto driver | 0.17 (0.1-0.31) | <0.001 | 0.42 (0.19-0.95) | 0.04 |
| Motor vehicle driver | 0.30 (0.08-1.13) | 0.08 | 0.25 (0.04-1.37) | 0.11 |
| Student | 1.11 (0.81-1.52) | 0.53 | 0.99 (0.52-1.87) | 0.97 |
| Housewife | 0.17 (0.12-0.25) | <0.001 | 1.01 (0.51-1.97) | 0.99 |
| Unemployed | 0.57 (0.34-0.96) | 0.04 | 0.67 (0.31-1.46) | 0.32 |
| Retied/Disabled/Sick | 0.65 (0.37-1.12) | 0.07 | 1.41 (0.52-3.82) | 0.5 |
| Other | 0.42 (0.17-1.06) | 0.07 | 1.08 (0.32-3.63) | 0.9 |
| Religion |  |  |  |  |
| Islam | Ref |  | Ref |  |
| Hinduism | 1.37 (0.86-2.20) | 0.19 | 1.30 (0.62-2.75) | 0.69 |
| Christianity | 6.86 (2.29-20.54) | 0.001 | 6.31 (0.996-40.01) | 0.05 |
| Region |  |  |  |  |
| Urban | Ref |  | Ref |  |
| Rural | 0.09 (0.07-0.12) | 0.01 | 0.12 (0.08-0.19) | <0.001 |
| Name of the vaccine |  |  |  |  |
| Sinopharm | Ref |  | Ref |  |
| OxfordAstraZeneca | 5.33 (3.56-7.98) | <0.001 | 4.51 (2.53-8.04) | <0.001 |
| Pfizer-BioNTech | 10.47 (6.03-18.19) | <0.001 | 5.37 (2.57-11.22) | <0.001 |
| Moderna | 7.1 (4.34-11.60) | <0.001 | 4.28 (2.28-8.05) | <0.001 |
| Sinovac | 0.68 (0.22-2.07) | 0.5 | 0.61 (0.15-2.50) | 0.5 |
| Don't know the name | 0.88 (0.51-1.51) | 0.64 | 1.62 (0.83-3.2) | 0.16 |
| Smoking status |  |  |  |  |
| No | Ref |  | Ref |  |
| Yes | 9.96 (7.47-13.26) | <0.001 | 3.6 (2.30-5.62) | <0.001 |
| Illicit substances |  |  |  |  |
| No | Ref |  | Ref |  |
| Yes | 5.13 (2.88-9.13) | <0.001 | 1.46 (0.61-3.48) | 0.4 |
| Diabetes |  |  |  |  |
| No | Ref |  | Ref |  |
| Yes | 2.12 (1.41-3.20) | <0.001 | 2.16 (1.15-4.06) | 0.02 |
| Hypertension/High blood pressure |  |  |  |  |
| No | Ref |  | Ref |  |
| Yes | 0.6 (0.65-1.01) | 0.06 | 0.32 (0.13-0.78) | 0.01 |
| Low blood pressure |  |  |  |  |
| No | Ref |  | Ref |  |
| Yes | 2.69 (1.63-4.44) | <0.001 | 3.33 (1.53-7.26) | 0.002 |
| Obesity |  |  |  |  |
| No | Ref |  | Ref |  |
| Yes | 2.9 (1.42-5.92) | 0.003 | 1.31 (0.44-3.91) | 0.63 |
| Severe allergic problems |  |  |  |  |
| No | Ref |  | Ref |  |
| Yes | 5.23 (2.89-9.46) | <0.001 | 4.17 (1.66-10.49) | 0.002 |
| Chronic respiratory diseases (Pneumonia, Asthma, breathing issues) |  |  |  |  |
| No | Ref |  | Ref |  |
| Yes | 2.91 (1.66-5.09) | <0.001 | 3.10 (1.32-7.30) | 0.01 |
| Anemia |  |  |  |  |
| No | Ref |  | Ref |  |
| Yes | 7.54 (2.55-22.32) | <0.001 | 4.61 (1.11-19.16) | 0.04 |

*UOR: Unadjusted Odds Ratio; AOR: Adjusted Odds Ratio; Ref: Reference group; CI: Confidence Interval*

The odds of experiencing COVID-19 vaccine side effects among female participants were 92% (95% CI: 0.11-0.32) lower than their male counterparts. Those aged 50-59 years and 60 or over were respectively 2.55 (95% CI: 1.04-6.24) times and 5.47(95% CI: 2.14-13.97) times more likely to experience side effects compared to the age group of 12 to 29 years. In comparison with the respondents with no formal education, those who had passed HSC (12^th^ grade), undergraduate, and graduate studies were less likely to experience side effects—71% (95% CI: 0.28-1.6), 66% (95% CI: 0.12-0.68) and 68% (95% CI: 0.14-0.81), respectively. The odds of experiencing side effects among rural respondents were 88% lower than their urban counterparts.

Smokers were 3.6 (95% IC: 2.30-5.62) times more likely to suffer from side effects than non-smoker respondents. Respondents who took illicit substances were 1.46 (0.61-3.48) times more likely to experience the COVID-19 vaccine’s side effects than those who did not (not statistically significant at 5% level). For underlying health conditions: those with low blood pressure displayed 3.33 (95% CI: 1.53-7.26) times higher chance to experience side effects; obese individuals were 1.31 (CI 0.44-3.91) times more likely; those suffering from severe allergies were 4.17 (95% CI: 1.66– 10.49) times more likely; those suffering from chronic respiratory diseases were 3.10 (95% CI: 1.32-7.30) times more likely; those suffering from anemia were 4.6 (CI 1.11-19.16) more likely than the participants with no underlying conditions.

### 3.4 Perception and Attitude Towards COVID-19 and Vaccination

Perception and attitudes towards COVID-19 and vaccinations are shown in Table 5. Most respondents either agreed that vaccines check against serious illness (50.59%) or remained neutral (42.09%). In addition, a majority agreed that all eligible people should take COVID-19 vaccines (72.14%) and maintain safety protocols even after vaccination (85.02%). Moreover, 77.78% of people agreed that the government and policymakers should make it mandatory for all eligible people to receive a COVID-19 vaccine.

**Table 5:** Perception and attitude towards COVID-19 vaccination

| Question | Agree | Neutral | Disagree |
| --- | --- | --- | --- |
| Covid-19 vaccines can protect you from serious COVID-19 illness (hospitalization, oxygen, ventilators, or death) | 50.59 (601) | 500 (42.09) | 7.32 (87) |
| All eligible people should take Covid-19 vaccines | 72.14 (857) | 24.75 (294) | 3.11 (37) |
| Even after getting fully vaccinated, we should maintain safety protocols (wearing mask, washing hands, avoiding gatherings, etc) | 85.02 (1,010) | 13.97 (166) | 1.01 (12) |
| Govt. and policymakers should make it mandatory for all eligible people to take a Covid-19 vaccine | 77.78 (924) | 20.03 (238) | 2.19 (26) |
| People should have a preference in choosing which Covid vaccine to take | 67.00 (796) | 27.36 (325) | 5.64 (67) |
|  | Extremely likely | Somewhat likely | Not at all likely |
| If available, how likely are you to allow children of your family (5 or older) to take Covid vaccines? | 25.59 (304) | 48.15 (572) | 26.26 (312) |
| If available, how likely are you to allow older people of your family (70 or older) to take Covid vaccines? | 40.57 (482) | 42.51 (505) | 16.92 (201) |
| How likely are you to wear a mask when you are outside/in public transport/shops/public places? | 55.72 (662) | 34.43 (409) | 9.85 (117) |
| How likely are you to recommend getting the COVID-19 vaccine to others? | 50.93 (605) | 37.63 (447) | 11.45 (136) |
| How likely is it that Covid-19 spreads all over Bangladesh again? | 19.19 (228) | 67.85 (806) | 12.96 (154) |

A considerable hesitancy was observed among the participants in allowing their children (5 years or older) to receive a Covid-19 vaccine. Only 25.59% of the respondents were extremely likely to let their children receive a Covid-19 vaccine when available to them. Furthermore, only 40.57% of the participants were found extremely likely to allow their older people (70 years or over) to take a COVID-19 vaccine. Most respondents chose not to take a stance on the likelihood of COVID-19 spreading across Bangladesh again (67.85%).

## 4 Discussion

The study investigated the side effects of all the COVID-19 vaccines being deployed in Bangladesh. About two-thirds of the 1,180 participants were males, and two-thirds were aged 50 years or older. Our study participants are relatively older, probably because COVID-19 vaccines were offered to older people on a priority basis in Bangladesh. However, there was almost a perfect balance in the proportions of urban and rural participants. The majority of the participants received the Sinopharm vaccine (66.5%).

The study revealed that less than half of the participants (39.48%) experienced at least one side effect after receiving a COVID-19 vaccine in Bangladesh. The side effects reported were regular and mild. The most-reported side effects were injection-site pain, fever, headache, redness/swelling at the injection site, and lethargy. The side effects existed on an average of 1-3 days only, and no instance of serious effects/hospitalization was found among the study participants. These findings are consistent with similar studies conducted in the Czech Republic, India and Saudi Arabia [20,26,27], although the Indian study reported a somewhat higher prevalence of side effects.

Side effects were more prevalent among those who received Pfizer-BioNTech and Moderna vaccines (about 80%), followed by the OxfordAstraZeneca vaccine (Figure 4). In contrast, the prevalence of side effects was substantially lower among those who received China-based Sinopharm and Sinovac vaccines (21%-28%). A study among health professionals in Slovakia found that after taking the mRNA-based COVID-19 vaccine, BNT162b2 (Pfizer), the great majority (91.6%) of Slovak health professionals experienced at least one side effect, which is persistent with our study. Furthermore, more than 70% of those who experienced side effects from Pfizer and Moderna vaccinations had to take medication. In contrast, only one-tenth of those who received the Sinopharm vaccine and experienced side effects had to take medication. The findings imply that mRNA-based Moderna and Pfizer vaccines cause stronger side effects than other vaccines.

Our study found a significant association between side effects and type of vaccines using the Sinopharm vaccine as the reference vaccine to compare. OxfordAstraZeneca, Pfizer-BioNTech, and Moderna vaccines showed respectively 4.51 (95% CI: 2.53-8.04) times, 5.37 (95% CI: 2.57-11.22) times, and 4.28 (95% CI: 2.28-8.05) times higher likelihood of causing side effects compared to the Sinopharm vaccine. Besides, women were less likely to report side effects following vaccination than their male counterparts. This is a mixed finding, with most studies reporting higher side effects among males [25–29] and others reporting the opposite [28]. Moreover, older people (>50 years) were more likely to report vaccine side effects than the younger ones, which also disagrees with most other studies [20,26,29]. The prevalence of side effects among rural participants was considerably lower than the urban participants. This might be attributed to the fact that most rural people received the Sinopharm vaccine, and we found that side effects were rare among those who received the Sinopharm vaccine.

Smokers exhibited a 3.6 (95% CI: 2.30-5.62) times higher likelihood of reporting side effects than non-smokers. In addition, those who had underlying health conditions (low blood pressure, severe allergic problems, chronic respiratory diseases, and anemia) showed a 3-4 times higher prevalence of side effects. Riad et al., in their study conducted among Slovak healthcare workers, also found a higher prevalence of side effects among people with underlying health conditions. However, the severity of side effects experienced by the people with underlying medical conditions was not any different in our study. Hence, people with underlying medical conditions should not hesitate to take a COVID-19 vaccine. Instead, they should take it immediately since they are at a higher risk for COVID-19 [30].

A lack of confidence about the efficacy of the vaccines was observed among participants. Only half of the respondents agreed to the statement “COVID-19 vaccines can protect you from serious COVID-19 illness (hospitalization, oxygen, ventilators, or death)”; others remained neutral or disagreed. Also, considerable hesitancy was found among the respondents in allowing children and older people to take a COVID-19 vaccine. Only one-fourth of the participants were ready to let their kids (five years or over) receive COVID-19 vaccines, while less than half of them were willing to allow their senior citizens (70 years or over). These findings are consistent with a survey conducted in the USA in October 2021. Only about one-third of parents of children aged 5 to 11 years (27%) were ready to acquire a vaccine for their younger child as soon as one is approved, while a third said they would wait to see how the vaccine worked [31].

Vaccines’ successes cannot be determined by only their side effects. A higher prevalence of minor side effects does not imply that a vaccine is inferior in function to another vaccine with a lower prevalence of side effects. The possibility of minor side effects following COVID-19 vaccination can be viewed positively: as a necessary precursor to a successful immunological response [32]. Vaccine side effects are almost always moderate and temporary, indicating that the vaccine is accomplishing its purpose of increasing IFN production, the body’s natural immune stimulant [32]. This study and many other studies conducted across the world found COVID-19 vaccines’ side effects regular and temporary [20,33–40]. Also, it is proven that COVID-19 vaccines effectively prevent serious COVID-19 illness (hospitalization, oxygen, ventilators, or death) [41]. Therefore, vaccines are the most powerful weapon available to us in the fight against the ever-pervasive COVID-19 pandemic.

### 4.1 Strengths and Limitations of the Study

To the best of the authors’ knowledge, this study is the first to investigate the potential side effects of several (five) COVID-19 vaccines. In addition, the study identified influential factors for experiencing side effects and their severity among the general people of Bangladesh. Furthermore, participants of this study were the general people. Most of the previous studies of this nature were conducted among healthcare workers only.

However, there are some limitations of this study. First, due to convenience sampling selection approaches that were part of the online survey approach, there might be some selection biases, such as fewer low education or illiterate participants. Second, since the study was online, voluntary, and self-administered, we cannot confirm the seriousness of all participants while filling out the questionnaire.

## 5 Conclusion

Less than half of the 1,180 participants (39.48%) reported at least one side effect after taking a COVID-19 vaccine in Bangladesh. The most reported side effects were injection site pain, fever, headache, redness/swelling at the injection site, and lethargy and were mild that lasted 1-3 days. Side effects were most prevalent among those receiving Pfizer-BioNTech and Moderna vaccines (approximately 80%) and were least prevalent among those receiving the China-based Sinopharm and Sinovac vaccines (21%-28%). Also, Pfizer-BioNTech and Moderna vaccines caused comparatively stronger side effects than the other vaccines.

OxfordAstraZeneca, Pfizer-BioNTech, and Moderna vaccines showed respectively 4.51 (95% CI: 2.53-8.04) times, 5.37 (95% CI: 2.57-11.22) times, and 4.28 (95% CI: 2.28-8.05) times higher likelihood of causing side effects compared to the Sinopharm vaccine. Moreover, males, older (>50 years), urban people, smokers, people with underlying health conditions exhibited a significantly higher likelihood of reporting side effects after receiving COVID-19 vaccines. A lack of confidence in vaccines’ efficacy and a substantial level of hesitancy in allowing children (age five years or over) and senior citizens (70 years or over) to receive COVID-19 vaccines were observed.

The findings of this study will help counter misinformation about the safety of COVID-19 vaccines and thus combat vaccine hesitancy, particularly in Bangladesh and other lower-income countries.

## Supporting information

Questionnaire

STROBE

## Data Availability

All data produced are available online at https://osf.io/h6qwa/?view_only=e888b489ae3845ffb23014757f8528cd

## Acknowledgments

We are grateful to all who spent their valuable time participating in the survey voluntarily and sharing the link with others. We are also grateful to those who helped people (older and younger) conduct the survey who did not have access to the internet and smart devices. In addition, we are enormously thankful to the researchers who provided their ratings to evaluate and finalize the instrument.

## Author Contributions

All the authors contributed significantly to the preparation of the final manuscript. MM and SM conceptualized and designed the study. MM and SM also developed the instrument with input and feedback from all other authors. AUM, PH, AM, MTA, FFA, AI, and MMR helped with data collection and supervision, data cleaning, writing, and proofreading. SM was also responsible for data analysis. In addition, MM and SM wrote the first draft of the manuscript. Finally, MSR, HRK, and MI supervised the entire study (continuous feedback, editing, proofreading, etc.). The order of the authors’ list indicates the level of contribution for each author in the entire study.

## Data Availability

The data supporting this study’s findings are openly available at <u>Open Science Framework</u>.

## Supplemental

Questionnaire; STROBE Checklist

