## Supplementary material for "Side Effects of COVID-19 Vaccines and Perceptions about COVID-19 and Its Vaccines in Bangladesh": Questionnaire

| **Section A** | **Eligibility/Basic information** | | |
| --- | --- | --- | --- |
| **Serial No.** | **Question** | **Options** | **Remarks** |
| **Q1** | **Consent:** Assalamu Alaikum/Adab (Greetings).  I would like to request you to take part in a small survey. This study aims to explore the side effects of the COVID-19 vaccines in Bangladesh. The survey includes questions regarding your COVID-19 vaccination and its side effects (if any). It also includes some socio-demographic questions. I hope this information will help the Bangladesh government and policymakers to make a better vaccination strategy to fight off the COVID-19 pandemic. You will not be asked any personal or sensitive questions. You will not be asked your name/identity. The survey may take 4-5 minutes to complete. Please note that your participation in the survey is entirely voluntary. You can reject it anytime during the survey.  Do you consent to participate in the study? | 1. Yes 2. No | If Q1=2, then shows “Thank you for your time” and ends the survey. |
| Q2 | Gender | 1. Male 2. Female |  |
| Q3 | (**Eligibility Check**)  How old are you (in years)? | 1. Less than 12 years 2. In between 12 to 17 3. In between 18 to 29 3. In between 30 to 39 4. In between 40 to 49 5. In between 50 to 59 6. In between 60 to 69 7. 70 or above | If Q3=1 , then shows “Unfortunately you are not eligible to take part in this survey as you are under 12, please stop and submit it” |
| **Q4** | (**Eligibility Check**)  Have you taken any Covid-19 vaccine (at least one dose)? | 1. Yes 2. No |  |
| **Q5** | Reason for not taking a vaccine yet | 1. Did registration, not yet received 2. Did not register, willing to take 3. I don’t want to take a vaccine 4. I don’t know 5. Others (specify) | **If Q4=2** |
| **Q6** | Why don’t you want a Covid-19 vaccine? | 1. It is very difficult to manage a vaccine  2. Vaccines aren’t safe and effective  3. Vaccines can cause death  4. People get infected and die even after taking the vaccines  5.Covid-19 isn’t a serious disease, I don’t fear  6. Only God (Allah/Vogoban) can save us  7. I have no time to go to the Vaccine center  8. I don’t know  8. Other (specify) | **If Q4=2 &Q5=3** |
| **Q7** | Why didn’t you register for the Covid vaccine? | 1. I don’t know how to register  2. I don’t have access to computers/smartphones/internet  3. I don’t know what it requires to get a vaccine  4. I do not have my NID/BR card or lost the NID/BR  5. Other (specify) | **If Q4=2 & Q5=2** |
| **if Q4=2, show *“Thank you for your time, unfortunately, you are not eligible to take part in this survey, please stop and submit”,* otherwise continue** | | | |
| **Section B: Demographics** | | | |
| Q8 | Your marital status | 1. Single 2. Married 3. Widowed 4. Divorced 5. Other(specify) |  |
| Q9 | Where do you live? | 1. Urban area (City corp, municipality) 2. Rural area |  |
| Q10 | What is your highest completed/running level of education? | 1. No formal education (Illiterate) 2. Primary completed 3. Higher secondary (grade 6-10) 4. SSC or equivalent completed 5. HSC or equivalent passed 6. Undergraduate (Hon’s/MBBS/Degree/Technical or Vocational) 7. Graduate (Maters/PhD/Mphil) 8. Other (specify) |  |
| Q11 | What is your religion? | 1. Islam 2. Hinduism 3. Christianity 4. Buddhism 5. Other religion (specify) |  |
| Q12 | In which of the following categories would you place your monthly household income from all sources before tax and any other deductions? (in Taka) | 1. Under 10,000 2. 10,000-19,999 3. 20,000-29,999 4. 30,000–39,999 5. 40,000–49,999 6. 50,000–74,999 7. 75,000 or over 8. Don't know |  |
| Q13 | What is your occupational status? | 1. Service holder (govt/private/RMG) 2. Small business (less than 5 employees) 3. Large business (5 or more employees) 4. Day laborer/Rickshaw/Van/Auto driver 5. Motor vehicle driver 6. Student 7. Housewife 8. Unemployed 9. Retired/disabled/sick 10. Other (specify) |  |
| **Section C Vaccination** | | | |
| Q14 | Which Covid-19 vaccine did you take? | 1. OxfordAstraZeneca (Oxford vaccine) 2. Pfizer-BioNTech (Pfizer vaccine) 3. Moderna 4. Sinopharm 5. Sinovac 6. Johnson & Johnson 7. Sputnik V 8. Don’t know the name | If Q4=1, the survey will continue. Otherwise, it will stop after the demographic part. |
| Q15 | Who/which influenced you to take the vaccine? | 1. I willingly took it 2. My family members/relatives convinced me 3. It was compulsory for workplace/educational institution 4. Neighbor/village leader inspired me 5. Others (specify) 6. Don’t know |  |
| **Section D** | **Underlying health conditions of respondents** | | |
| Q.15 | Which of the following underlying conditions do you have? (Select all that apply) | 1. No underlying conditions 2. Diabetes 3. Heart disease/Heart attack 4. Hypertension/High blood pressure 5. Low blood pressure 6. Cancer 7. Obesity 8. Severe allergic problems 9. Chronic respiratory diseases (Pneumonia, Asthma,   breathing issues)   10. Liver/kidney disease 11. Anemia 12. Other regular health issues (specify) |  |
| Q16 | Do you smoke | 1. Yes  2. No |  |
| Q17 | Do you drink (Alcohol)/take illicit substances (Gaja/Yaba, etc.) | 1. Yes  2. No |  |
| **Section E** | **Side effects (if Q4=1)** | | |
| Q18 | Did you have any side effects after taking the vaccine (normal or severe)? | 1. Yes 2. No | If Q-18=1, then ask all the side-effects related questions |
| **Now we will ask you some questions about the side effects you faced after taking the covid vaccine** | | | |
| Q19 | Pain at the vaccination site | 1. Severe pain 2. Normal pain 3. No pain |  |
| Q19.1 | How many days did it last (no. of days) |  | If Q19=1 or 2 |
| Q20 | Redness/swelling at the vaccination site (ঈঞ্জেকশনের স্থানে লাল/ফুলা ভাব) | 1. Yes 2. No |  |
| Q20.1 | How many days did it last (no. of days) |  | If Q20=1 |
| Q21 | Fever | 1. Normal fever 2. Severe fever 3. No fever |  |
| Q21.1 | How many days did it last (no. of days) |  | If Q21=1 or 2 |
| Q22 | Headache | 1. Yes 2. No |  |
| Q22.1 | How many days did it last (no. of days) |  | If Q22=1 |
| Q23 | Lethargy/tiredness | 1. Yes 2. No |  |
| Q23.1 | How many days did it last (no. of days) |  | If Q23=1 |
| Q24 | Nausea | 1. Yes 2. No |  |
| Q24.1 | How many days did it last (no. of days) |  | If Q24=1 |
| Q25 | Diarrhea | 1. Yes 2. No |  |
| Q25.1 | How many days did it last (no. of days) |  | If Q25=1 |
| Q26 | Cough | 1. Yes 2. No |  |
| Q26.1 | How many days did it last (no. of days) |  | If Q26=1 |
| Q27 | Allergic reaction | 1. Yes 2. No |  |
| Q27.1 | How many days did it last (no. of days) |  | If Q27=1 |
| Q28 | Muscle pain | 1. Yes 2. No |  |
| Q28.1 | How many days did it last (no. of days) |  | If Q28=1 |
| Q29 | Abdominal Pain | 1. Yes 2. No |  |
| Q29.1 | How many days did it last (no. of days) |  | If Q29=1 |
| Q30 | Did you feel anxious after taking the vaccine? | 1. Yes  2. No |  |
| Q31 | Did you experience a decrease in sleep quality? | 1. Yes  2. No  3. Don’t know |  |
| Q32 | Did you experience an increase in sleep time? | 1. Yes  2. No  3. Don’t know |  |
| Q33 | Did you take any medicine (s) due to the side effects? | 1. Yes 2. No |  |
| Q34 | How serious were the side effects of the vaccine? | 1. Not serious, I was able to do all my usual activities 2. I couldn’t do my usual activities for 1 day 3. I couldn’t do my usual activities for more than 1 day 4. Side effects were enough to get worried 5. I needed to get hospitalized/see a doctor 6. I don’t know |  |
| Q35 | How different were the reactions of the two doses? | 1. Yet to take the second dose 2. Reactions after the first dose were more severe 3. Reactions after the second dose were more severe 4. There was no difference 5. Don’t know |  |
| Q36 | Has anybody discouraged you from taking the 2^nd^ dose because of the side effect in your first dose? | 1. Yes 2. No |  |
| **Section F** | **Perceptions about Covid-19 and its vaccines** | | |
| Q37 | How safe do you think you are after getting vaccinated? | 1. 100% safe, will never get infected with Covid-19 2. Feel safe, but still, I could get infected 3. No confidence, I can get infected anytime 4. Don’t know | If Q4=1 |
| Q38 | Covid-19 vaccines can protect you from serious COVID-19 illness (hospitalization, oxygen, ventilators, or death) | 1. Agree 2. Neutral 3. Disagree |  |
| Q39 | All eligible people should take Covid-19 vaccines | 1. Agree 2. Neutral 3. Disagree |  |
| Q40 | Even after getting fully vaccinated, we should maintain safety protocols (wearing mask, washing hands, avoiding gatherings, etc) | 1. Agree 2. Neutral 3. Disagree |  |
| Q41 | Govt. and policymakers should make it mandatory for all eligible people to take a Covid-19 vaccine | 1. Agree 2. Neutral 3. Disagree |  |
| Q42 | Govt. and specialists should decide which Covid vaccine is suitable for whom (considering age, gender, health conditions, etc) | 1. Agree 2. Neutral 3. Disagree |  |
| Q43 | People should have a preference in choosing which Covid vaccine to take | 1. Agree 2. Neutral 3. Disagree |  |
| **N1** | If available, how likely are you to allow children of your family (5 or older) to take Covid vaccines? | Not at all likely  Somewhat likely  Extremely likely |  |
| N4 | If available, how likely are you to allow older people of your family (70 or older) to take Covid vaccines? | Not at all likely  Somewhat likely  Extremely likely |  |
| N6 | How likely are you to wear a mask when you are outside/in public transport/shops/public places? | Not at all likely  Somewhat likely  Extremely likely |  |
| N7 | How likely are you to recom­mend getting the COVID-19 vaccine to others? | Not at all likely  Somewhat likely  Extremely likely |  |
| N8 | How concerned are you about getting COVID-19? | Not at all concerned  A little concerned  Moderately concerned  Very concerned |  |
| N9 | How do you feel about the amount of information on COVID-19 and its vaccines that you are getting? | I’m not getting  enough information.  I’m getting  enough information.  I’m getting too  much information. |  |
| N10 | What are your sources of information about Covid-19 and its vaccines (select all that apply)? | Social media (facebook/instagram/twitter, etc)  TV/Radio/Newspaper  Family/Friends  Neighbors/Other people  Local health officials/Covid websites  WHO/CDC/FDA  Others (specify) |  |
| N11 | How difficult it is for general people to get Covid vaccines in Bangladesh? | Very easy  Somewhat easy  Somewhat difficult  Very difficult  Not sure |  |
| N12 | How likely is it that Covid-19 spreads all over Bangladesh again? | Not at all likely  Somewhat likely  Extremely likely |  |
| Q44 | Do you have any idea about the cost of vaccines that the government is spending | 1.Yes  2.No |  |
| Q45 | How much cost for each dose |  | If Q 44 is 1 |
